# Comparison of antigen- and RT-PCR-based testing strategies for detection of Sars-Cov-2 in two high-exposure settings

**DOI:** 10.1101/2021.06.03.21258248

**Authors:** Jay Love, Megan T. Wimmer, Damon J.A. Toth, Arthi Chandran, Dilip Makhija, Charles K. Cooper, Matthew H. Samore, Lindsay T. Keegan

## Abstract

Surveillance testing for infectious disease is an important tool to combat disease transmission at the population level. During the SARS-CoV-2 pandemic, RT-PCR tests have been considered the gold standard due to their high sensitivity and specificity. However, RT-PCR tests for SARS-CoV-2 have been shown to return positive results when administered to individuals who are past the infectious stage of the disease. Meanwhile, antigen-based tests are often treated as a less accurate substitute for RT-PCR, however, new evidence suggests they may better reflect infectiousness. Consequently, the two test types may each be most optimally deployed in different settings. Here, we present an epidemiological model with surveillance testing and coordinated isolation in two congregate living settings (a nursing home and a university dormitory system) that considers test metrics with respect to viral culture, a proxy for infectiousness. Simulations show that antigen-based surveillance testing coupled with isolation greatly reduces disease burden and carries a lower economic cost than RT-PCR-based strategies. Antigen and RT-PCR tests perform different functions toward the goal of reducing infectious disease burden and should be used accordingly.

## Introduction

Since its emergence in late 2019, SARS-CoV-2, the virus responsible for COVID-19, has spread rapidly, causing significant global morbidity and mortality. Although early outbreaks were concentrated in China and Italy, the United States (US) was the global epicenter for most of 2020, accounting for approximately one fourth of all cases globally by March 2021 (Dong, Du, & Gardner, 2020). Despite the approval of multiple SARS-CoV-2 vaccines, production, distribution, and uptake hurdles combined with the emergence of novel viral variants indicates that achieving herd immunity remains a distant prospect (Moore & Offit, 2021; Sallam, 2021). Therefore, comprehensive testing, contact tracing, and infectious case isolation remain important interventions to continue slowing the spread of SARS-CoV-2 and maintaining health system integrity (Love et al., 2021).

As testing availability has increased, epidemiological questions have arisen regarding the optimal deployment of different test strategies. Diagnostic tests for SARS-CoV-2, none of which perfectly reflect viral carriage (Woloshin et al. 2020), fall into two broad categories: antigen tests and real-time reverse transcription polymerase chain reaction (RT-PCR) tests. While both tests diagnose active SARS-CoV-2 infection, antigen tests detect the presence of a specific viral antigen and are capable of returning results within 15 minutes, while RT-PCR amplify genomic sequences and therefore require longer turn-around times (CDC 2021a). Substantial attention has been paid to the lower sensitivity of antigen testing compared with that of RT-PCR testing (Scohy et al., 2020). Early studies on SARS-CoV-2 antigen testing sensitivity reported relatively low sensitivity with respect to RT-PCR, leading some public health officials to place lower confidence in antigen testing than RT-PCR testing in ending the COVID-19 pandemic. However, it has been suggested that when comparing antigen- and RT-PCR tests to viral culture, a proxy for transmissibility, one rapid antigen test (Becton Dickinson Veritor) had a negative percent agreement of 96.4% (95% CI: 82.3, 99.4) and a positive percent agreement of 90.0% (CI: 76.3, 97.6), while positive percent agreement for the RT-PCR assay was 73.7% (CI: 60.8, 85.3) (Pekosz et al., 2021). These studies suggest that antigen tests may perform better than RT-PCR tests at detecting actively infectious infections, since RT-PCR tests may detect low levels of viral nucleic acid that may not indicate current infectiousness (CDC 2020a). Consequently, the two different test types may have different optimal strategies for deployment. The more rapid antigen tests may be more optimally deployed when a delay in test results will delay the isolation of an infectious infection (e.g., Larremore et al., 2021), while RT-PCR tests may be more optimally deployed when diagnosing every infection or disease case is critical (e.g., Wang et al., 2020).

Recent studies have demonstrated that regardless of test sensitivity, widespread, high-frequency testing is sufficient to significantly reduce the burden of COVID-19 (Larremore et al., 2021; Mina, Parker, & Larremore, 2020; Paltiel, Zheng, & Walensky, 2020). While this finding highlights the importance of surveillance testing, it does not provide an explicit decision-support framework for clinical and public health decisionmakers for testing strategies in congregate settings. Building on such past work, we investigate the population-level epidemiological effects of employing different COVID-19 surveillance testing strategies, including RT-PCR, antigen, and two reflex testing strategies, in two non-acute care congregate living settings that bookend the risk spectrum: a nursing home and a university residence hall system.

## Methods

### Epidemiological model

The epidemiological model is comprised of a dynamical model of SARS-CoV-2 with a layered statistical model of hospitalizations, ICU hospitalizations, and deaths.

#### Transmission dynamics model

We developed a compartmental model of SARS-CoV-2 transmission dynamics with surveillance testing by modifying a *Susceptible-Exposed-Infectious-Removed* (SEIR) model and including individual-based accounting of infectious state (Figure 1). There are two simultaneous processes in the model: the disease process and the testing process. In the disease process (Figure 1), individuals start as fully susceptible (*S*) and become exposed (*E*) according to a density-dependent probability of exposure. This probability is defined as the product of a rate *β* and the infectious proportion of the population, accounting for quarantine and isolation, as described below (see Equation S1). The value of *β* is derived from the product of R_0_, which we assume to be uniformly distributed between 1.2 and 1.5 (Zhang, Keegan, Qiu, & Samore, 2020), and the recovery rate, *γ*, which we assume to be uniformly distributed between 1/2.6 and 1/6 per day (Lin et al., 2020). Exposed individuals have had a successful exposure event but are not yet infectious; they become infectious (*I*) at a rate *σ* and are then able to infect others. We assume *σ* is 1/3 (Peirlinck, Linka, Sahli Costabal, & Kuhl, 2020). Infected individuals are removed/recover (*R*) at a rate *γ*. The removed/recovered compartment indicates individuals who are no longer infectious, which includes both recovered and dead individuals; we use the classic epidemiologic definition of this term, and do not use it to refer to a clinical recovery. We seed our simulations based on a prevalence of 1.8% in the residence hall setting and 3.6% in the nursing home setting.

**Figure 1.**
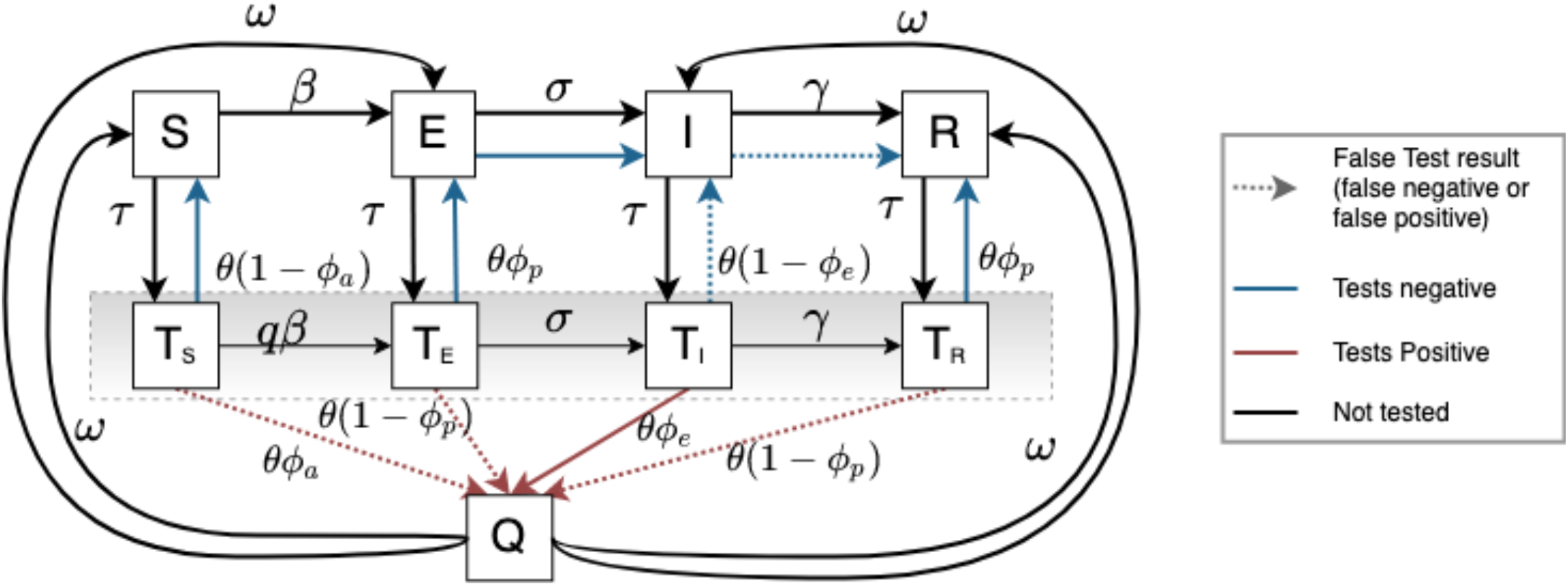
Schematic of the COVID-19 epidemiologic model. Individuals in the population start as susceptible (*S*) and become exposed (*E*) at a rate *β*. Exposed individuals become infectious (*I*) at a rate *σ*, and recover (R) at a rate *γ*. Simultaneously, surveillance testing occurs at a rate proportional to the population make up, and individuals awaiting test results are in a “leaky” quarantine. Tested susceptibles (*T*_*S*_) can become exposed (*T*_*E*_), then infectious (*T*_*I*_), and can infect susceptibles (both *S* and *T*_*S*_). Since tested infectious individuals (*T*_*I*_) are quarantining, their infectiousness is reduced by a factor *q*. Likewise, since tested susceptibles (*T*_*S*_), are also quarantining, their susceptibility is reduced by a factor *q*. Individuals who test positive are isolated (Q). We assume isolation is perfect, and thus individuals can only progress through their disease process (*Q*_*E*_ -> *Q*_*I*_ -> *Q*_*R*_); susceptibles who are isolated cannot become infected and infected individuals who are isolated cannot cause infections. After the isolation period is over (14 days in the standard condition), individuals are returned to the general population, retaining their current disease state. Antigen and RT-PCR tests have a specified positive (*ϕ*_*e*_) and negative percent agreement (*ϕ*_*p*_) with viral culture (see text). *ϕ*_*a*_ is 0.0001, representing imperfect test specificity. Note that we use positive percent agreement for tests of infectious individuals (*I*) and negative percent agreement for both exposed (*E*) and recovered (*R*) individuals, as it has been shown that RT-PCR may detect infection among individuals who are no longer infectious (Kohmer et al., 2021). Here, the color and line-type of the arrows indicate whether or not the test was “correct” or “incorrect” with respect to infectiousness, with dotted lines indicating incorrect test results (i.e., false negative or false positive), red lines indicating a positive test, blue lines indicating a negative test, black lines indicating that no test result was returned.

In the testing process (Figure 1), we assume a fixed proportion of the population, τ, is tested each day (surveillance testing). The assumptions and parameter values for our baseline simulation settings are as follow (Table S1). We assume that susceptible individuals who test positive (*T*_*S*;_ false positive) do so at a rate of *ϕ*_*a*_= 0.0001 to account for imperfect test specificity. For tested exposed (*T*_*E*_) and tested recovered (*T*_*R*_) individuals, we use the negative percent agreement of each test compared to culture reported in published analyses for RT-PCR (*ϕ*_*p*_= 95.5%) and for antigen tests (*ϕ*_*p*_= 98.7%) (Pekosz et al., 2021). For tested infectious individuals (*T*_*I*_), we use the antigen test positive percent agreement *ϕ*_*e*_= 96.4% and RT-PCR positive percent agreement *ϕ*_*e*_= 100% reported in the same analysis (Pekosz et al., 2021). We assume that individuals who test positive are isolated (*Q*) and are returned from isolation after an average of 14 days (*ω* = 0.07 in figure 1, though return from isolation is determined as 1/ω days after the start of isolation; see below); we also explore a shorter 10-day isolation (see supplement). We assume that patients will reduce their mixing while awaiting test results and therefore are both less susceptible and less infectious. As a baseline assumption, we use a 50% reduction in mixing due to this partial quarantine (*q*). We also explore two additional possible reductions of 25% and 75% (Figures S6, S7). Other than this partial quarantine, tested individuals move through the disease course as normal. Test results for antigen testing are returned on the same day as test administration (θ = 1), while results are returned in 48 hours for RT-PCR (θ = 1/3), an approximation for the US average time for RT-PCR test result turnaround (as in Larremore et al., 2021). We explore two additional possible RT-PCR test turnaround times of 1 and 4 days in the supplement.

We assume isolation resulting from a positive test result is perfect, and thus individuals can only progress through their disease process (*Q*_*E*_ -> *Q*_*I*_ -> *Q*_*R*_); susceptibles who are isolated cannot become infected and infected individuals who are isolated cannot cause infections. After the isolation period is over (14 days in the standard condition), individuals are returned to the general population, retaining their current disease state.

#### Test Settings

We selected two settings to bookend the risk spectrum for communal living scenarios: a nursing home, where disease outcomes are more severe, and a university residence hall system, where disease outcomes are less severe. In the nursing home setting, the population was 101, hospitalization rate was 25% of infections, ICU admission rate was 35.3% of hospitalizations, and the fatality rate was 5.4% of total infections. In the residence hall system setting, the population was 3150, hospitalization rate was 3.9% of infections, ICU admission rate was 23.8% of hospitalizations, and the fatality rate was 0.01% of total infections (values reflect estimates from CDC pandemic planning scenarios and expert guidance; CDC 2020b).

#### Testing strategies

We explore five testing strategies for each of these two settings and assumed asymptomatic screening of 1%, 2%, 5%, and 10% of the population each day. In each setting and for each testing strategy, we ran the simulation for a duration of 183 days. The five testing strategies are as follows:

1. Test once with RT-PCR test (“stand-alone PCR”)
2. Test once with antigen test (“stand-alone antigen”)
3. Test once with antigen test, then retest all negative results with antigen test two days later (“reflex to antigen”)
4. Test once with antigen test, then retest all symptomatic negative results with RT-PCR test (“reflex symptomatic to PCR”)
5. No surveillance testing (“no testing”)

Testing strategy 4 relies on symptom presentation to flag symptomatic individuals with negative antigen tests for retesting. We assume that 60% of all infections are symptomatic (CDC 2020b). To account for the potential influence of other circulating respiratory infections, we also assume a background rate of 2% of the population expressing symptoms of influenza-like illness that are unrelated to but could be confused with infection with SARS-CoV-2 (Tokarz et al., 2019; CDC 2021b). Individuals expressing influenza-like illness symptoms are randomly drawn from the population each day with a 2% probability. Individuals expressing symptoms of COVID-19 or influenza-like illness are flagged for retesting with RT-PCR in this strategy. Those individuals flagged for retesting are quarantined in the same way as individuals awaiting test results.

To determine the relative impact of different testing strategies on the disease course, we compare a suite of metrics between the four testing strategies and against the output of the model in the absence of testing. The metrics reported are number of infections averted, hospitalizations averted, deaths averted, and the per test reduction in infections.

Since using testing metrics with respect to viral culture as a proxy for infectiousness is a novel approach to modeling infectious disease surveillance testing strategy effectiveness, we also conducted simulations using antigen test sensitivity and specificity with respect to RT-PCR test results. RT-PCR is capable of detecting levels of viral RNA that do not indicate infectiousness (i.e., “recovered” in our model) (Mina et al., 2020). While our main simulations use test positive percent agreement to determine test results exclusively for infectious individuals, in these alternate simulations, we use test sensitivity to determine test results for both infectious and recovered individuals less than 54 days after recovery, approximating the median duration of RT-PCR detectable viral shedding in one analysis of 36 patients (Li, Wang, & Lv, 2020). We use test specificity to determine test results for susceptible and exposed classes and recovered individuals more than 54 days after recovery in these alternate simulations. Test sensitivity and specificity are positive percent agreement with RT-PCR results, considered the testing standard. Therefore, RT-PCR sensitivity and specificity are 100%, whereas antigen test sensitivity was 84.7% and specificity was 99.5%, following published results (Young et al. 2021).

We constructed the model in the R statistical environment (R Core Team, 2017). The discrete-time model includes flows between states that are determined by rates at the population level and by functions at the individual-level. The time step is one day. The probability of exposure is frequency dependent and varies at each time step and across individuals. It is the product of the infected and unquarantined proportion of the population at each time step and *β* plus the product of the infected and quarantined proportion, the mixing rate reduction due to quarantine (*q*), and *β. β* is determined by the product of random draws from uniformly-distributed values of *R0* and *γ*. The probability of becoming infectious is determined by the rate *σ*. Recovery, however, is modeled not with a population-level rate (*γ*), but with an individual-based approach using a fixed duration of infectiousness; for each infection, a recovery day is designated that is 1/*γ* time steps from the day of infection. Similarly, tests are returned by defining a day of test return for each test that is 1/θ time steps from the day of testing, and return from isolation is determined by defining an end day to the isolation period as 1/ω days after isolation begins.

We conducted 10,000 stochastic simulations for each testing strategy in each setting.

### Economic Model

To improve the decision support aspect of our model, we layered a cost effectiveness analysis on top of the epidemiological model to estimate the costs of each testing strategy relative to the relevant outcomes. We used parameter values estimated from literature sources and expert input (Table S2). All costs were expressed in 2021 US dollars and are from the perspective of the congregate setting decision makers. We estimated total testing and outcome costs per strategy and calculated Cost per Incremental Infection Avoided and Unnecessary Quarantine Costs. We also calculated Incremental Cost Effectiveness Ratios (ICER) to compare the four testing strategies with the no testing strategy using the following standard calculation: 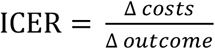.

#### Testing Strategy Costs

We considered the direct cost of testing to be the product of the number of tests per day, price per test per day, personal protective equipment (PPE) costs per day, and labor costs per day. For each type of test, the total cost is the sum of these daily costs multiplied by the duration of the simulation. We assume that RT-PCR is sent to an external laboratory and therefore has no direct capital costs to the decision maker, and we assume PPE is the same for either type of testing. The no test strategy by definition has no direct testing costs.

#### Outcomes Cost Model

We then consider the direct costs to the decision makers who purchase and conduct testing in each of our two settings: universities and nursing homes. For nursing homes, staff absenteeism due to quarantine incurs costs measured in labor productivity loss, a cost limited to staff, and a conservative proxy for true costs which could include temporary staff and other costs. Positive test results in either residents or staff incur labor costs for the administrative burden of reporting positives and initiating cleaning protocols. For positive residents, additional PPE and labor costs are incurred due to assumed need for isolation and additional staff care for those residents. Total outcomes cost is therefore calculated as:

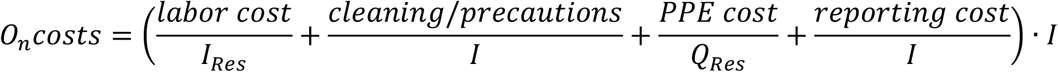

where

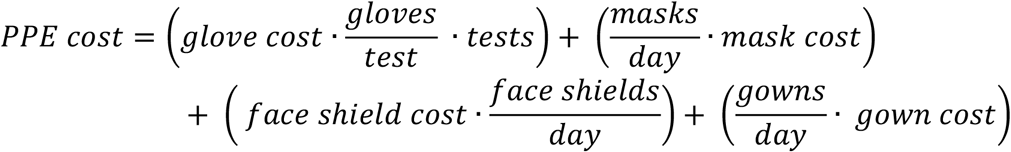

and where labor cost is the product of the hourly wage and the number of hours worked for each healthcare worker type.

For university residence halls, the cost per infection (*O*_*n*_*costs*) was provided based on expert input.

See Table S2 for cost parameter values and references.

## Results

Compared to simulations without testing, all testing strategies reduced the peak and total infections in simulated epidemics (Figure 1). Greater reduction in infections was achieved with higher rates of daily screening. The relative differences between testing strategies’ performance in reducing infections were largely maintained across both nursing home and dormitory settings.

In the nursing home setting, no statistically significant differences were found in % infections averted across testing strategies at low levels of surveillance testing (1%, 2%). At high levels of surveillance testing (5%, 10%), the reflex to antigen strategy (34% and 53% averted) outperformed standalone PCR (26% and 44% averted), which outperformed standalone antigen (20% and 35% averted) (Figures 2,3). The reflex symptomatic negatives to PCR strategy did not statistically significantly improve performance versus standalone antigen in any case (Figure 2).

**Figure 2.**
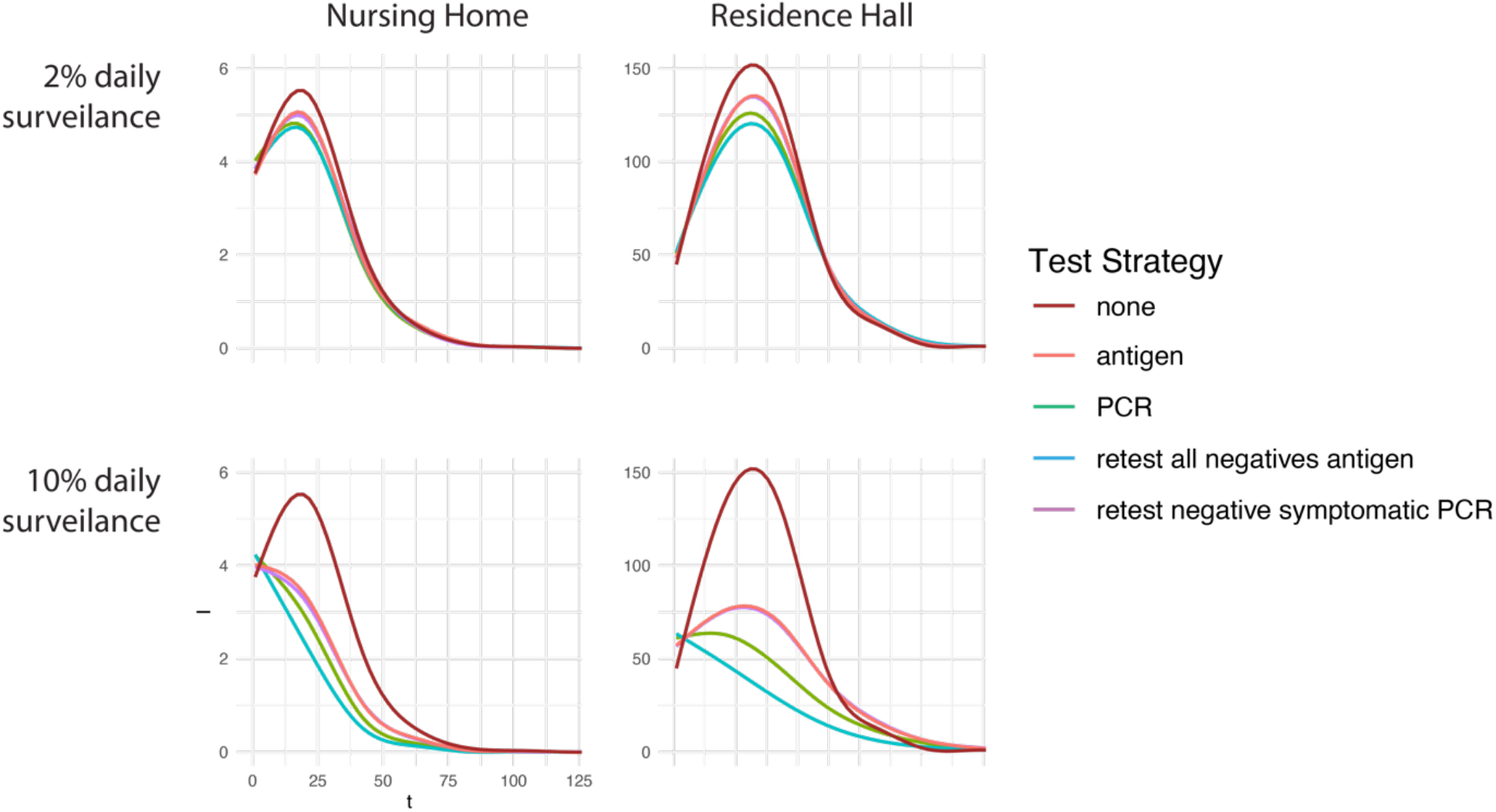
Epidemic curves showing infections over time in the nursing home (left) and residence hall (right) group living settings and at 2% and 10% daily surveillance testing. Higher levels of testing reduce infections. Different test strategies perform similarly. Line color indicates testing strategy.

**Figure 3.**
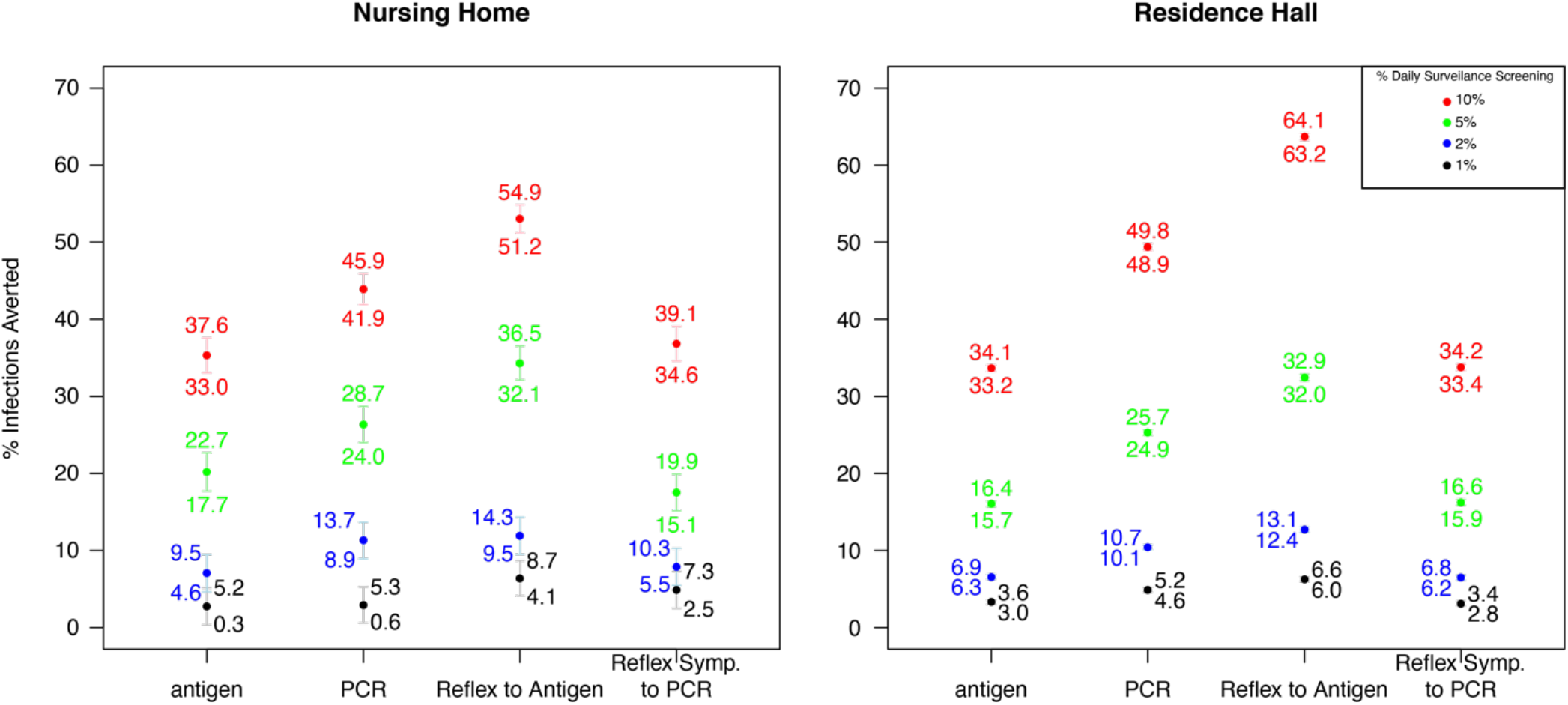
Percent infections averted for different testing strategies at different daily surveillance testing percentages. Red = 10%, green = 5%, blue = 2%, black = 1% daily surveillance testing. Filled circles show means, whiskers show 95% CI. Text labels indicate values of upper and lower 95% confidence interval bounds.

These same broad trends were also observed in the university residence hall setting. For university dormitories under screening rates of 1%, the reflex to antigen strategy averted the highest percentage of infections (∼6%), followed by standalone PCR (∼5%), then standalone antigen (∼3%). Standalone antigen testing and reflexing symptomatic to PCR strategies were again statistically equivalent in infection reduction (∼3%) (Figure 2). At higher daily screening rates, this overall pattern was largely conserved, yet exaggerated. For 5% and 10% daily testing, overall reduction of infection rates increased to 16-32%, and 34-64%, respectively (Figure 2).

Since hospitalizations and deaths are calculated as proportions of infections, hospitalizations averted and deaths averted follow patterns similar to that found in infections averted (Figure S1). Very few deaths occurred in the residence hall setting.

On a per-test basis, different testing strategies resulted in similar percent infections averted (Figure S8). At higher surveillance levels (i.e., 5% and 10% daily surveillance) and with larger populations (i.e., in the residence hall setting), standalone PCR testing showed a notably higher mean per-test percent infection averted (Figure S8).

Overall, surveillance testing reduces the disease burden in populations, and the effect of testing on measures of disease burden was greatest at the highest rates of testing. We found similar patterns in simulations that used different parameter values for days to PCR test return, quarantine mixing reduction, and days of isolation (See Figures S2-S7).

When evaluating antigen testing by modeling test sensitivity and specificity with respect to PCR, we find that the performance difference between standalone antigen and PCR testing is greater than when these test features are modeled with respect to viral culture (Figure S2). For example, the difference between standalone PCR and antigen testing strategies in mean % infections averted in the residence hall setting at 10% daily surveillance testing was roughly 20% when evaluating test performance with respect to PCR (Figure S2b) but only roughly 15% when evaluating test performance with respect to viral culture (Figure 3).

The economic analysis revealed that antigen-based strategies carried an overall lower economic cost than did RT-PCR-based strategies. At 2% and 10% screening rates, respectively, over the 6-month period in nursing homes, the testing strategy of standalone antigen testing was least expensive in terms of overall testing costs overall testing costs ($25,037 and $48,445), total costs ($38,250 and $77,460) and testing cost per infection ($544 and $1,514) (Table 1). For university dormitories, a similar pattern was evident (testing costs $486,074 and $952,940, total costs $798,147 and $1,445,972, testing cost per infection $419 and $840, at 5% and 10% screening per 6 months, respectively) (Table 1). The overall testing and total costs are higher for the university residence hall setting, reflecting the larger population and higher number of tests needed. In the residence hall setting, the cost per person is lower than that of the nursing home setting, demonstrating efficiencies of scale.

**Table 1.** Results of the economic cost analysis. In both settings and at all levels of surveillance testing, the antigen-based testing strategy was least expensive. The “retest all negatives with antigen” strategy, which averted the most infections (Figure 3), was less expensive than PCR-based testing. The “retest negative symptomatic with PCR” strategy was similar in costs to the antigen-based testing strategy.

| Surveillance % | Setting |  | Testing strategy | Testing cost per infection | Total testing cost | Total Costs | Per person testing cost | Cost per incremental infection avoided |
| --- | --- | --- | --- | --- | --- | --- | --- | --- |
| 1% | NH | 1 | PCR | \$659.40 | \$31,652.69 | \$43,324.77 | \$10.05 | \$12,840.77 |
| | NH | 2 | Antigen | \$459.99 | \$22,125.35 | \$29,174.74 | \$7.02 | \$9,007.33 |
| | NH | 3 | retest all negatives with antigen | \$539.16 | \$24,959.52 | \$35,762.94 | \$7.92 | \$6,012.60 |
| | NH | 4 | retest negative symptomatic PCR | \$471.29 | \$22,167.77 | \$29,708.94 | \$7.04 | \$6,707.82 |
|  | NH | 5 | None |  |  |  |  |  |
| | Dorms | 1 | PCR | \$298.35 | \$485,242.81 | \$609,273.18 | \$154.05 | \$4,043.76 |
| | Dorms | 2 | Antigen | \$68.10 | \$112,601.12 | \$184,479.47 | \$35.75 | \$1,485.94 |
| | Dorms | 3 | retest all negatives with antigen | \$126.32 | \$202,452.15 | \$348,270.93 | \$64.27 | \$1,714.37 |
| | Dorms | 4 | retest negative symptomatic PCR | \$67.97 | \$112,599.40 | \$185,112.30 | \$35.75 | \$1,521.70 |
|  | Dorms | 5 | None |  |  |  |  |  |
| 2% | NH | 1 | PCR | \$1,063.56 | \$46,654.07 | \$67,441.43 | \$14.81 | \$7,461.16 |
| | NH | 2 | antigen | \$544.68 | \$25,037.09 | \$38,250.02 | \$7.95 | \$5,331.76 |
| | NH | 3 | retest all negatives with antigen | \$703.20 | \$30,636.99 | \$49,059.50 | \$9.73 | \$4,538.34 |
| | NH | 4 | retest negative symptomatic PCR | \$551.15 | \$25,109.82 | \$38,597.40 | \$7.97 | \$5,058.64 |
|  | NH | 5 | None |  |  |  |  |  |
| | Dorms | 1 | PCR | \$622.28 | \$953,359.76 | \$1,182,796.20 | \$302.65 | \$3,926.41 |
| | Dorms | 2 | antigen | \$128.87 | \$205,871.92 | \$344,489.52 | \$65.36 | \$1,432.23 |
| | Dorms | 3 | retest all negatives with antigen | \$257.34 | \$384,030.53 | \$652,701.97 | \$121.91 | \$1,667.43 |
| | Dorms | 4 | retest negative symptomatic PCR | \$128.84 | \$206,013.34 | \$345,351.85 | \$65.40 | \$1,440.51 |
|  | Dorms | 5 | None |  |  |  |  |  |
| 5% | NH | 1 | PCR | \$3,066.44 | \$91,330.75 | \$129,235.64 | \$913.31 | \$6,569.19 |
| | NH | 2 | antigen | \$1,063.42 | \$33,839.19 | \$59,210.23 | \$338.39 | \$3,357.35 |
| | NH | 3 | retest all negatives with antigen | \$1,987.85 | \$47,376.39 | \$76,359.32 | \$473.76 | \$2,979.99 |
| | NH | 4 | retest negative symptomatic PCR | \$1,030.46 | \$33,973.28 | \$60,181.84 | \$339.73 | \$3,650.04 |
|  | NH | 5 | none |  |  |  |  |  |
| | Dorms | 1 | PCR | \$2,263.99 | \$2,359,441.85 | \$2,808,400.54 | \$749.03 | \$4,205.48 |
| | Dorms | 2 | antigen | \$418.97 | \$486,074.53 | \$798,147.93 | \$154.31 | \$1,451.72 |
| | Dorms | 3 | retest all negatives with antigen | \$1,088.13 | \$919,568.27 | \$1,425,698.88 | \$291.93 | \$1,648.46 |
| | Dorms | 4 | retest negative symptomatic PCR | \$419.71 | \$486,327.46 | \$797,692.80 | \$154.39 | \$1,447.13 |
|  | Dorms | 5 | none |  |  |  |  |  |
| **10%** | NH | 1 | PCR | \$5,981.02 | \$165,931.49 | \$210,071.39 | \$1,659.31 | \$6,880.82 |
| | NH | 2 | antigen | \$1,514.33 | \$48,445.07 | \$77,460.72 | \$484.45 | \$2,695.88 |
| | NH | 3 | retest all negatives with antigen | \$3,199.94 | \$74,309.03 | \$102,585.67 | \$743.09 | \$2,777.84 |
| | NH | 4 | retest negative symptomatic PCR | \$1,556.56 | \$48,636.12 | \$77,438.30 | \$486.36 | \$2,636.02 |
|  | NH | 5 | none |  |  |  |  |  |
| | Dorms | 1 | PCR | \$5,427.84 | \$4,700,756.49 | \$5,260,632.24 | \$1,492.30 | \$4,680.53 |
| | Dorms | 2 | antigen | \$839.73 | \$952,940.34 | \$1,445,972.64 | \$302.52 | \$1,484.75 |
| | Dorms | 3 | retest all negatives with antigen | \$2,864.65 | \$1,779,042.18 | \$2,312,562.13 | \$564.78 | \$1,677.51 |
| | Dorms | 4 | retest negative symptomatic PCR | \$842.05 | \$953,376.96 | \$1,444,826.84 | \$302.66 | \$1,480.44 |
|  | Dorms | 5 | none |  |  |  |  |  |

Due to the low number of occurring retests, the reflex symptomatic to PCR strategy had costs very similar to standalone antigen testing (Table 1). Importantly, stand-alone PCR resulted in the highest costs of all of the testing strategies.

Strategies that resulted in longer test result wait time increased unnecessary quarantine costs, with stand-alone PCR resulting in the highest of these across settings at all screening rates, ranging from 1.61-2.34 times as costly as the testing strategy with the least unnecessary quarantine costs (Table 1). ICERs were calculated for all testing strategies. Antigen-based strategies averaged lower ICERs than PCR-based strategies (Table 1).

## Discussion

Our model demonstrates that surveillance testing paired with quarantine and isolation is likely to reduce disease burden in congregate living settings during the COVID-19 pandemic. Higher rates of screening result in the strongest reduction in infections and are associated with greater distinction between the performance of different testing strategies. At low screening rates, testing strategies performed similarly to each other. Despite differences in population size and underlying risk, both residence hall and nursing home settings showed reduced disease burden with increased surveillance testing.

Notably, the degree to which the standalone PCR testing strategy prevents infections is highly contingent on the effectiveness of quarantine during the test-result waiting period. At low waiting-period quarantine effectiveness, standalone PCR testing outperforms standalone antigen testing to a lesser degree than at higher waiting-period quarantine effectiveness (Figures 2, 3, S6, S7). If individuals awaiting surveillance test results do not undergo quarantine, as may occur in some settings, the difference between these two testing strategies is likely to less pronounced.

While the reflex to antigen strategy was the most effective at reducing infections, standalone antigen testing was the least expensive and most cost-effective testing strategy in both settings at all screening rates, due primarily to differences in test prices. Standalone PCR testing performed well, but it was outperformed by the reflex to antigen strategy, both in terms of economic cost and of reducing disease burden.

During the COVID-19 pandemic, RT-PCR testing has been established as the standard by which to measure other tests. Our modeling analysis demonstrates that using viral culture, which may better reflect viral transmissibility (Pekosz et al., 2021), as the test standard dramatically alters the relative performance of different surveillance testing strategies. Under this paradigm, antigen-based surveillance testing strategies coupled with infectious case isolation are shown to strongly reduce disease burden at a level close to RT-PCR-based strategies, but at a much lower economic cost (Figures 2,3), somewhat in contrast to model results under a more typical RT-PCR test-standard paradigm (Figure S2). This lower cost has the potential to make additional resources available for other management, containment, or recovery efforts, and therefore has the potential to substantially reduce disease burden during the COVID-19 pandemic or others in the future. Understanding test results with the priority endpoint in mind (e.g., the test’s ability to identify currently infectious infections during surveillance testing programs) should be of primary importance, and our modelling study should prompt further research on the relationship between viral culture, diagnostic test results, and transmissibility for SARS-CoV-2 and other infectious diseases.

The cost perspective of this model may be of use to public health decision makers in determining whether or not to invest in surveillance testing, but it does not account for the broader costs to society and the healthcare system. An expanded or alternative perspective to this model that could estimate the indirect societal costs of infection, disease, and quarantine would likely yield more robust cost-effectiveness values and ICERs compared to those we find here.

Our results support the work of other studies that demonstrate that frequency of testing can overcome differences in sensitivity (e.g., Larremore et al. 2021). In an important addition, we provide a pragmatic example of an affordable and effective strategy that is implementable in two group-living settings. As vaccine uptake remains low in many group living settings (e.g., Cavanaugh et al. 2021), and as a greater understanding of the potential for immune escape mutants is developed (Garcia-Beltran et al., 2021), surveillance testing strategies that use antigen tests can be considered as highly effective, cost-reducing alternatives to PCR testing strategies.

## Data Availability

Modeling study - no empirical data used

