## Supplement for "Comparison of antigen- and RT-PCR-based testing strategies for detection of Sars-Cov-2 in two high-exposure settings"

Equation S1.

$$P_{expose} = \frac{\beta(I_n + qI_a)}{N}$$

$I_n$ = # unisolated infectious individuals not awaiting test results
$I_a$ = # unisolated infectious individuals awaiting test results
$q$ = reduction in mixing rate due to partially-effective quarantine while awaiting test results
$N$ = population size.

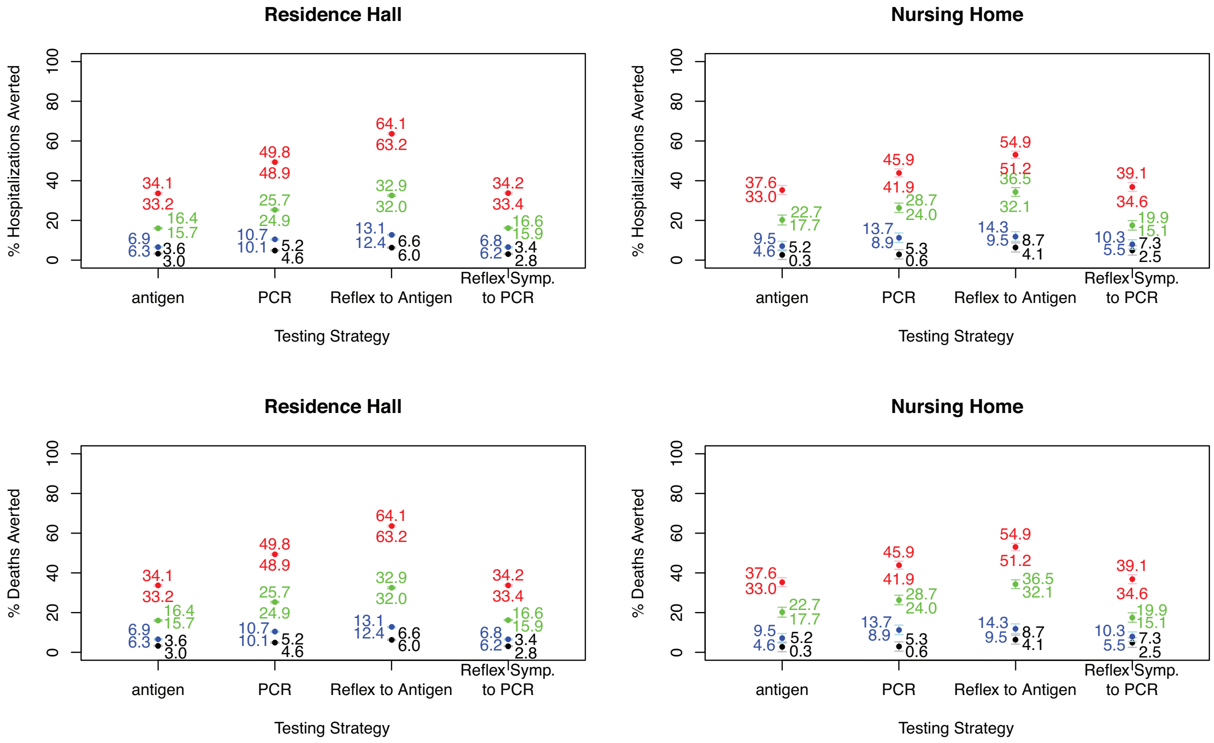

Figure S1. Percent deaths and percent hospitalizations averted. Since hospitalizations and deaths are calculated as proportions of infections, hospitalizations averted and deaths averted follow patterns similar to that found in infections averted (Figure 2).

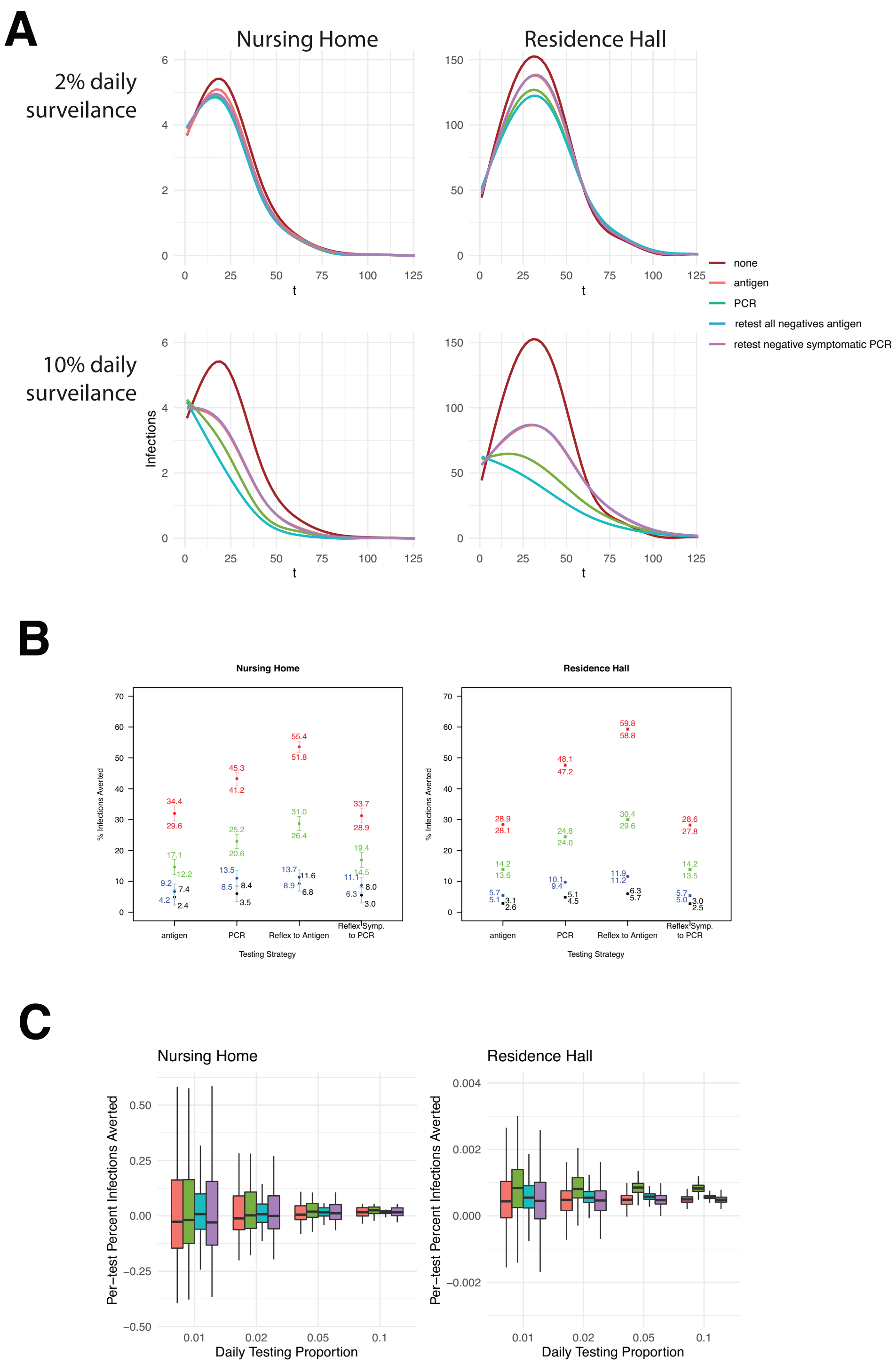

Figure S2. Model results when evaluating antigen test performance with respect to PCR. In our main results (Figures 2,3), we evaluate test performance with respect to viral culture. When evaluating performance with respect to PCR, the test standard for COVID-19 diagnostic testing, a standalone RT-PCR-based test strategy outperforms a standalone antigen-based strategy by a larger margin than when evaluating performance with respect to viral culture (see Figures 2,3 in the text).

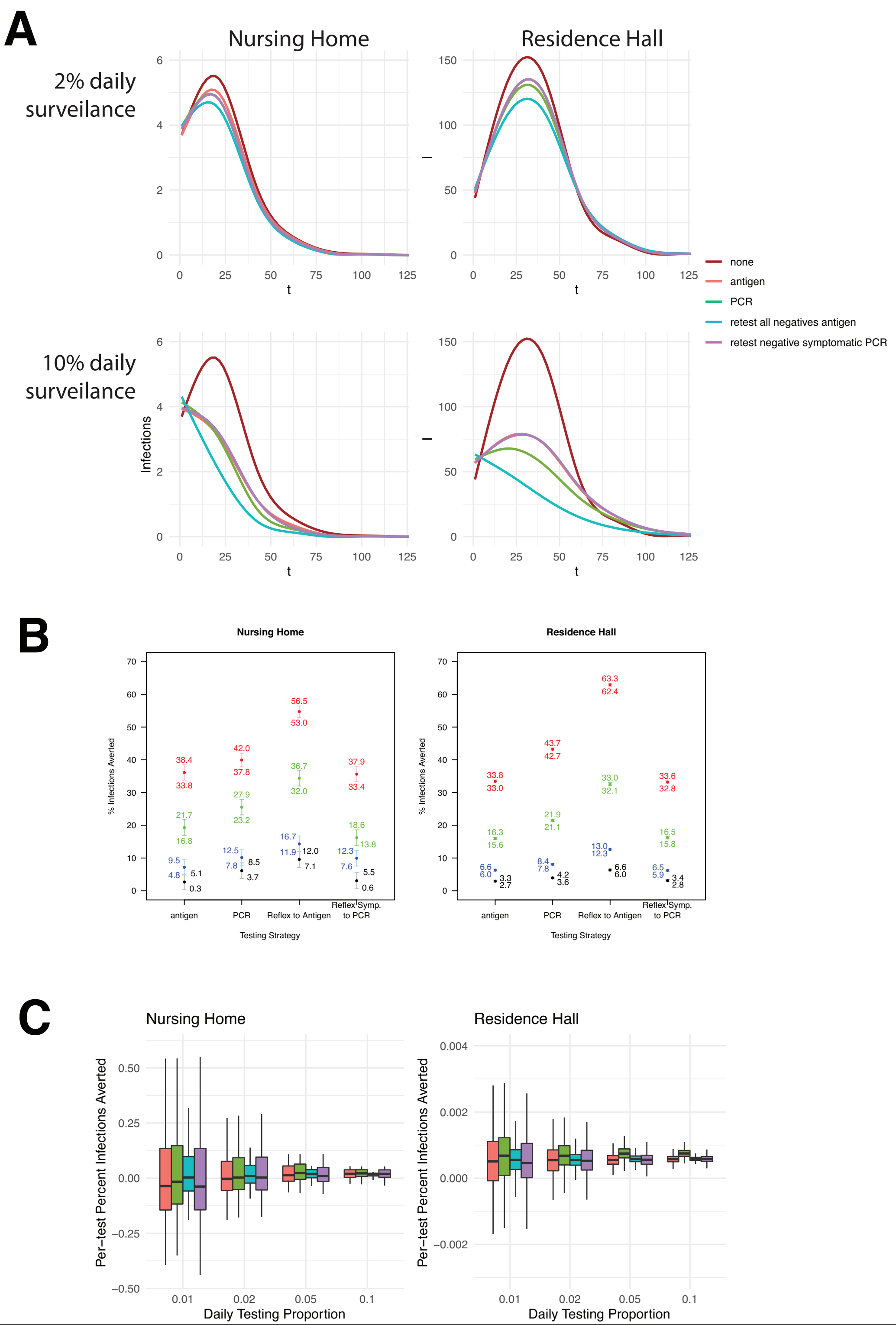

Figure S3. Model results when RT-PCR test result turnaround time is 1 day. Patterns are similar to those in the main results, which used a longer turnaround time, but in this case the PCR-based strategy averts fewer infections than in the main results. This is likely attributable to reduced time spent in partial quarantine while awaiting test results.

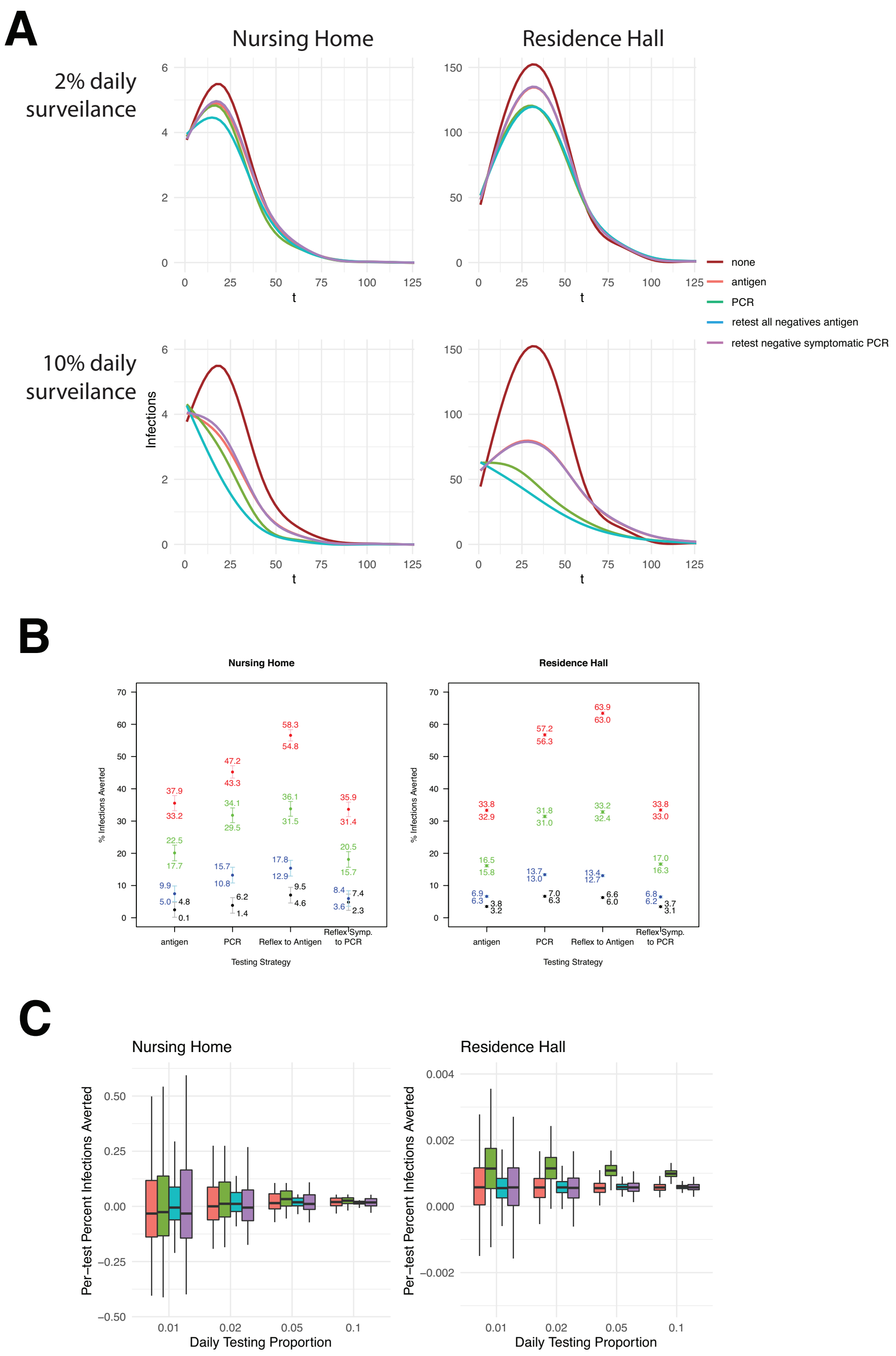

Figure S4. Model results when RT-PCR test result turnaround time is 4 days. Patterns are similar to those in the main results, which used a shorter turnaround time, but in this case the PCR-based strategy averts more infections than in the main results. This is likely attributable to increased time spent in partial quarantine while awaiting test results.

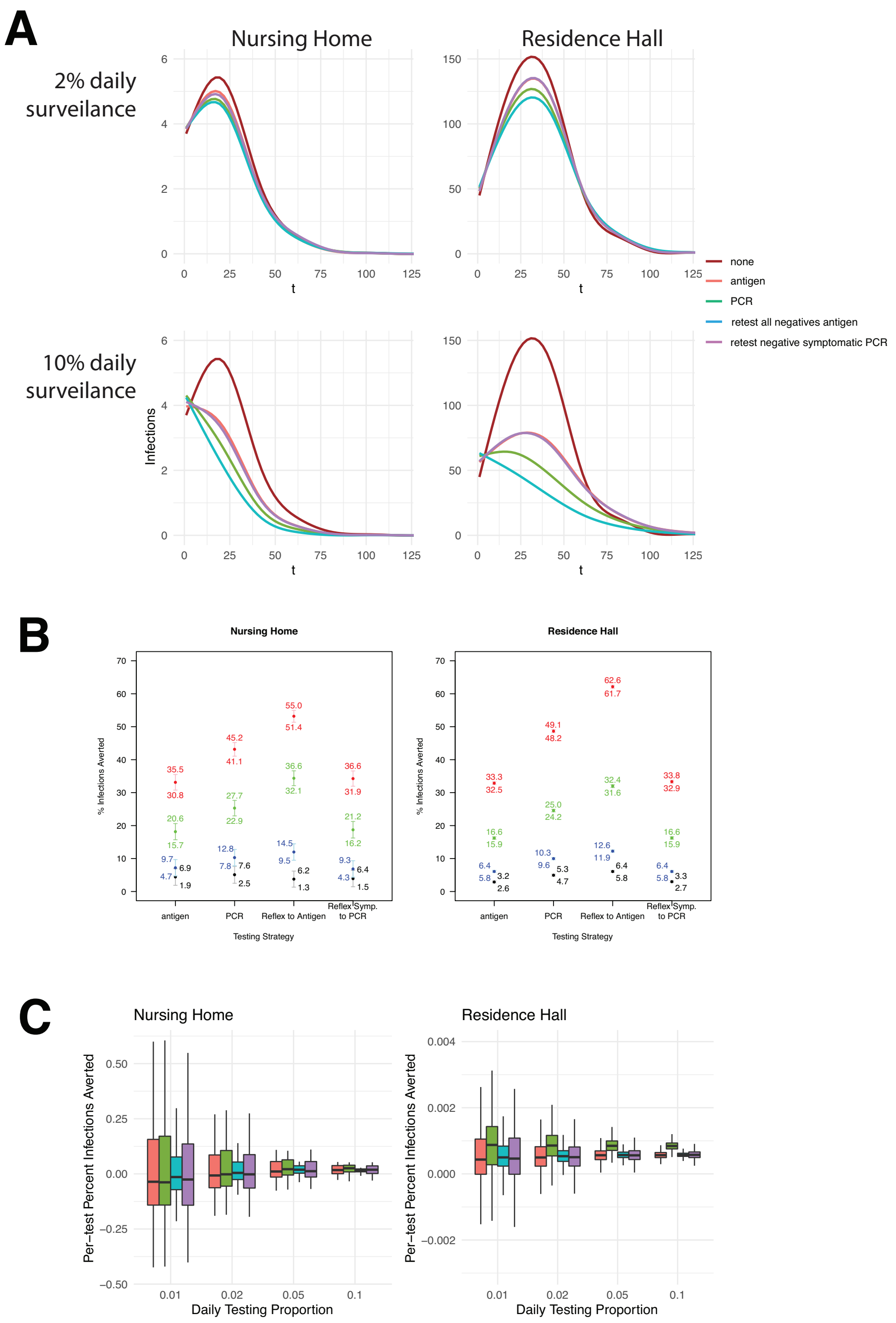

Figure S5. Model results when the test-positive isolation period is 10 days. Patterns are very similar to those in the main results, which used a 14 day isolation period.

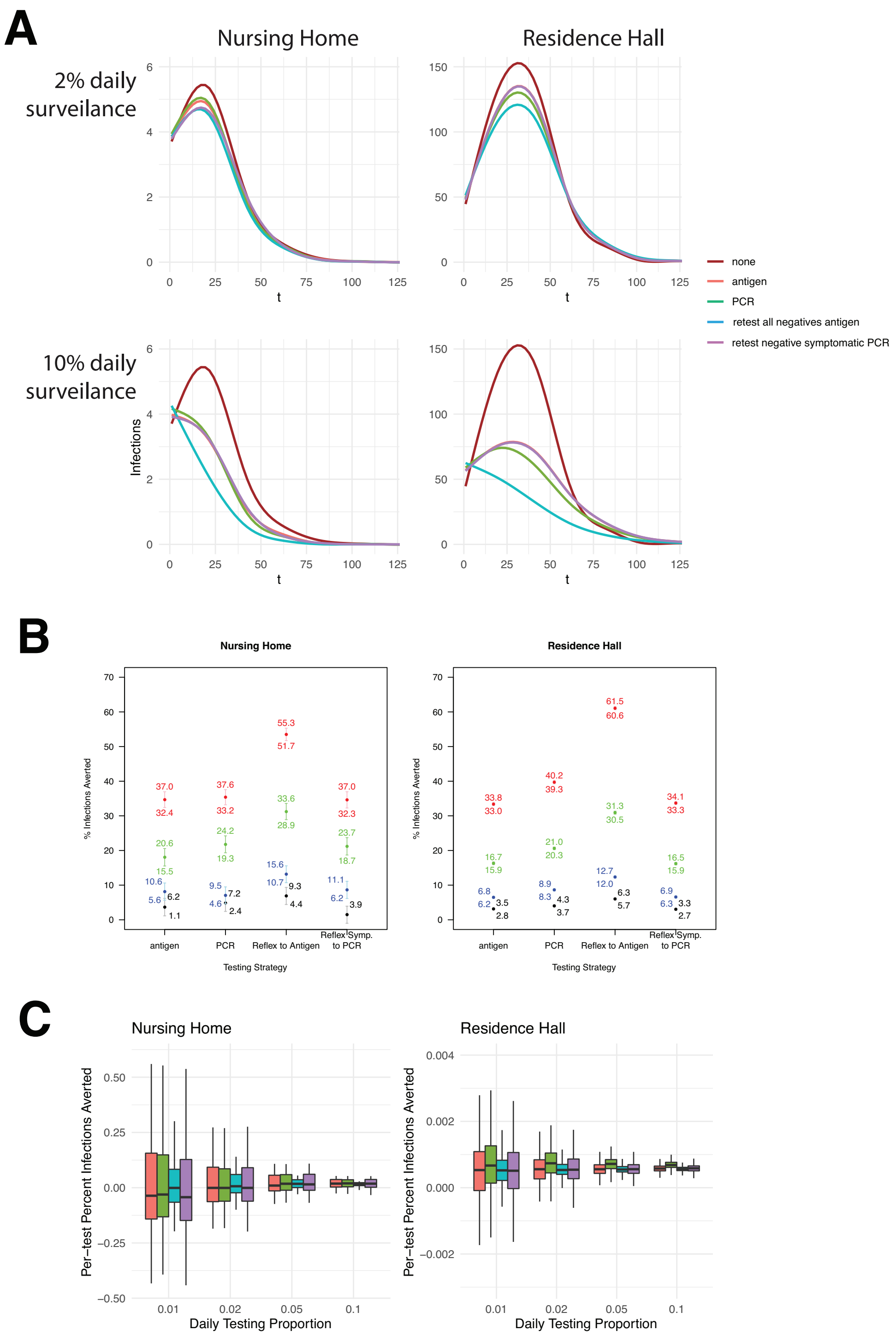

Figure S6. Model results when the mixing reduction during quarantine while waiting for test results is 25%. Patterns are similar to those in the main results, which used a reduction of 50%, but in this case fewer infections are averted in the PCR-based strategy, where waiting times were longest.

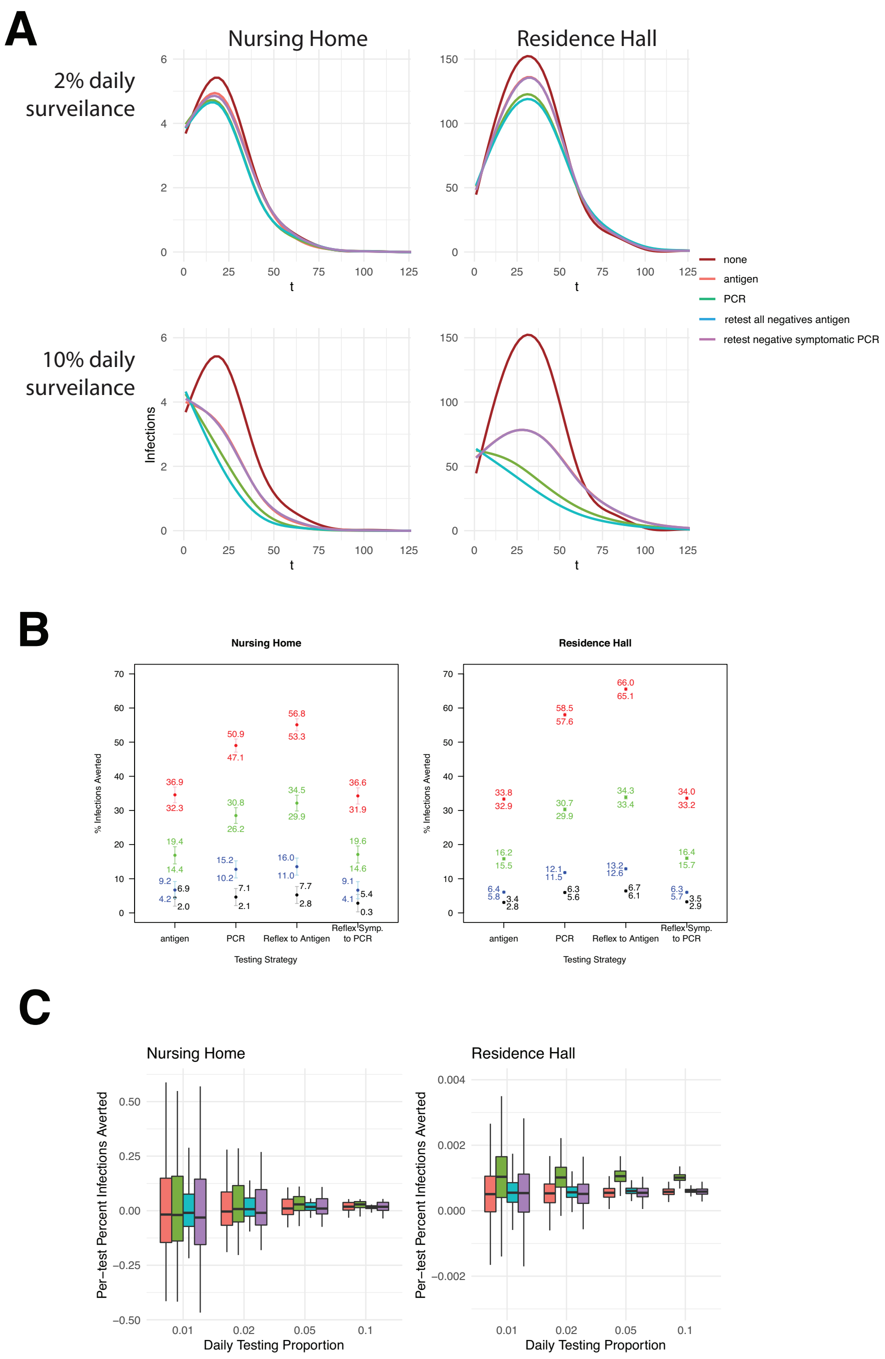

Figure S7. Model results when the mixing reduction during quarantine while waiting for test results is 75%. Patterns are similar to those in the main results, which used a reduction of 50% but in this case more infections are averted in the PCR-based strategy, where waiting times were longest.

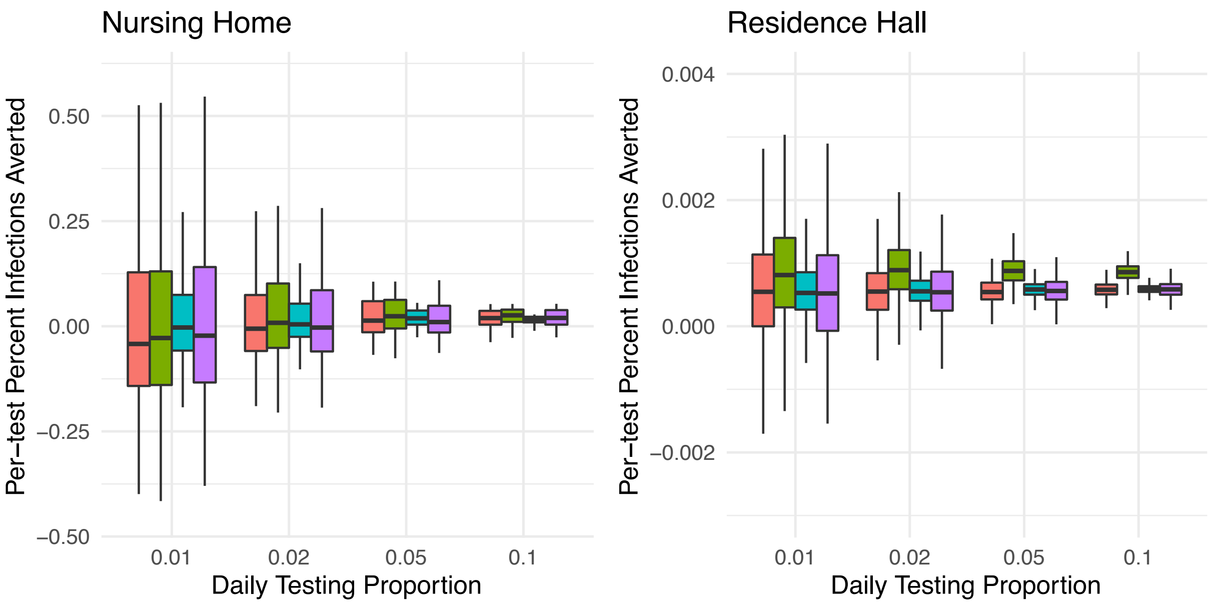

Figure S8. Per-test percent infections averted for different testing strategies at different daily surveillance testing percentages. Colors indicate different test strategies, as in Figure 1. Boxes show median, 1st, and 3rd quartiles and whiskers show extent of data up to 1.5 times the interquartile range.

Table S1. Test parameter values for the baseline simulation settings.

| | RT-PCR | antigen |
|---|---|---|
| $\phi_a$ | 0.01% | 0.01% |
| $\phi_p$ | 95.50% | 98.70% |
| $\phi_e$ | 100% | 96.40% |
| θ | 0.33 | 1 |

Table S1

| Nursing Home | Value | Units | Min | Max | Source (*All 2020 dollars adjusted to 2021) |
|---|---|---|---|---|---|
| Number Tests Run Per Day | **521.52** | # tests | | | |
| Infection Duration, Days | **14** | days | | | |
| ***Cleaning Due to Outbreak*** | | | | | |
| Hours Spent Per Infection on Cleaning | **3.75** | hours | 1 | 5 | https://www.fourth.com/blog/managing-increased-cleaning-costs-in-a-covid-19-operation/ |
| Staff Hourly Wage, Nursing Home, Cleaning | **$ 14.02** | USD | $12.69 | $14.41 | http://www.nahc.org/wp-content/uploads/2017/10/10hc_stats.pdf |
| ***Caring for Residents in Quarantine*** | | | | | |
| Staff Hourly Wage, Nursing Home, Patient Care | **$ 18.16** | USD | $13.88 | $ 22.88 | http://www.nahc.org/wp-content/uploads/2017/10/10hc_stats.pdf |
| HCW Labor Hours Spent per Quarantined Resident Per Day | **3.8** | hours | 2.57 | 7 | https://www.cdc.gov/nchs/data/series/sr_03/sr03_43-508.pdf, |
| PPE Costs Per Care of Quarantined Resident per Day | **$ 94.15** | USD | $44.37 | $177.49 | https://www.nheconomy.com/getmedia/e1c2a698-e7a1-4e18-8391-b66b52c0bd38/Average_PPE_Price.pdf |
| ***Labor Force Loss (HCW only) Due to Quarantine*** | | | | | |
| Hours of Productivity per HCW lost per day per infection | **12** | hours | 8 | 12 | assumption |
| Staff HCW Hourly Wage | **$ 18.16** | USD | $13.88 | $22.88 | http://www.nahc.org/wp-content/uploads/2017/10/10hc_stats.pdf |
| ***Contact Tracing and Reporting Costs*** | | | | | |
| Hours Spent Per Infection on Contact Tracing and Reporting | **0.5875** | hours | 0.375 | 0.8 | https://www.cdc.gov/coronavirus/2019-ncov/php/contact-tracing/COVIDTracer.html |
| Staff Hourly Wage, Nursing Home, Reporting/ Contact Tracing | **$ 20.56** | USD | $11.50 | $29.94 | https://www.indeed.com/career/contact-tracer/salaries |
| ***Testing Labor Costs*** | | | | | |
| Hours Spent Per Test Collection & Send Out, PCR | **0.067** | hours | n/a | | BD internal data |
| Hours Spent Per Test Run/Collection, Antigen | **0.050** | hours | | | |
| Staff Hourly Wage, Nursing Home, Running Test/ Test Collection | **$ 18.16** | USD | $13.88 | $ 22.88 | |

| **Nursing Home** | | | | |
|---|---|---|---|---|
| *PPE Costs Per Test* | $91.41 | _ | _ | |
| Cost of a Gown | $2.72 | USD | 1 per shift breaks, 1 break per shift | https://www.nheconomy.com/getmedia/e1c2a698-e7a1-4e18-8391-b66b52c0bd38/Average_PPE_Price.pdf |
| Cost of a Mask (N95) /unit | $8.94 | USD | 2 per shift breaks, 1 break per shift | |
| Cost of Gloves | $0.05 | USD | 1 per shift patient + 1 extra | |
| Cost Per Papper | $11.04 | USD | 1 per shift | |
| Gloves Per Test | 2 | USD | 1 | assumption |
| Masks Per Day of Testing | 2 | unit | 1 | |
| Gowns per Day of Testing | 2 | unit | 1 | |
| Pappers Per Day Testing | 1 | unit | 1 | |
| *Capital Costs Veritor* | $2,476.00 | | No capital PCR testing costs due to sending out | BD internal data |
| Veritor Analyzers | $299.00 | 1 | | |
| NUC | $1,200.00 | 1 | *optional add on | BD internal data |
| Infoscan | $399.00 | 1 | *optional add on | |
| Barcoding printer | $275.00 | 1 | *optional add on | |

| | | | | |
|---|---|---|---|---|
| Synapsys Cost | $300.00 | 1 | *optional add on | |
| Timers | $3.00 | 1 | | |
| *Variable Per Test Costs* | | | | |
| Per Test Cost Veritor | $15.00 | | | BD internal data |
| Per Test Cost PCR (price send out) | $80.00 | | | |

| **University** | | | | |
|---|---|---|---|---|
| Hours Spent Per Infection on Cleaning | 3.75 | hours | 1 N95 mask, 1 gown, 1 face shield per shift, 2 pair gloves = per cleaning FTE per clean, 7.5 hour shift, 50% = 3.75 hours per day more cleaning. | https://www.fourth.com/blog/managing-increased-cleaning-costs-in-a-covid-19-operation/ |
| Staff Hourly Wage, University, Cleaning | $15.45 | USD | Janitor salary not special for COVID-19 | https://www.salary.com/research/salary/benchmark/janitor-sr-salary |
| Cost per positive (isolation per student) | $643.75 | USD | | Expert Input (Utah) |
| Hours Spent Per Infection on Contact Tracing and Reporting | 0.5875 | hours | assumes 10% of time that full contact tracing would take, reporting only, where contact tracing takes 3.75 to 8 hours per case per CDC datasheet | https://www.cdc.gov/coronavirus/2019-ncov/php/contact-tracing/COVIDTracer.html |
| Staff Hourly Wage, University, Reporting/ Contact Tracing | $20.56 | USD | A contact tracer is the next most relevant hourly salary | https://www.indeed.com/career/contact-tracer/salaries |
| Hours Spent Per Test Collection & Send Out, PCR | 0.067 | hours | BD time in motion data | BD internal data |
| Hours Spent Per Test Run/Collection, Antigen | 0.05 | hours | BD time in motion data | |
| Staff Hourly Wage, University, Running Test/ Test Collection | $25.75 | hours | | https://www.cnbc.com/2020/05/20/temperature-screeners-covid-19-testers-these-jobs-were-created-by-pandemic.html |
| *PPE Costs Per Test* | | | | |
| Cost of a Gown | $2.72 | USD | 1 per shift breaks, 1 break per shift | https://www.nheconomy.com/getmedia/e1c2a698-e7a1-4e18-8391-b66b52c0bd38/Average_PPE_Price.pdf |
| Cost of a Mask (N95) | $8.94 | USD | 2 per shift breaks, 1 break per shift | |
| Cost of Gloves | $0.05 | USD | 1 per shift patient + 1 extra | |
| Cost Per Papper | $11.04 | USD | 1 per shift | |
| Gloves Per Test | $2.00 | USD | 1 | assumption |
| Masks Per Day of Testing | 2 | unit | 1 | |
| Gowns per Day of Testing | 2 | unit | 1 | |
| Pappers Per Day Testing | 1 | unit | 1 | |
| *Capital Costs Veritor* | 0 | n/a | No capital PCR testing costs due to sending out | BD internal data |
| Veritor Analyzers | $299.00 | 1 | 0 | |
| NUC | 1,200.00 | 1 | *optional add on | |
| Infoscan | $399.00 | 1 | *optional add on | |
| Barcoding printer | $275.00 | 1 | *optional add on | |
| Synapsys Cost | $300.00 | 1 | *optional add on | |
| Timers | $3.00 | 1 | | |
| *Variable Per Test Costs* | | | | |
| Per Test Cost Veritor (price) | $15.00 | 1 | | BD internal data |
| Per Test Cost PCR (send out) | $80.00 | 1 | | |

| University | Value | Units | Min | Max | Source |
|---|---|---|---|---|---|
| ***Cleaning Due to Outbreak*** | | | | | |
| Hours Spent Per Infection on Cleaning | 3.75 | hours | 1 | 5 | https://www.fourth.com/blog/managing-increased-cleaning-costs-in-a-covid-19-operation/ |
| Staff Hourly Wage, University, Cleaning | $15.00 | USD | $13.00 | $17.00 | https://www.salary.com/research/salary/benchmark/janitor-sr-salary |
| ***Contact Tracing and Reporting Costs*** | | | | | |
| Hours Spent Per Infection on Contact Tracing and Reporting | 0.5875 | hours | 0.375 | 0.8 | https://www.cdc.gov/coronavirus/2019-ncov/php/contact-tracing/COVIDTracer.html |
| Staff Hourly Wage, University, Reporting/ Contact Tracing | $19.96 | USD | $11.50 | $29.94 | https://www.indeed.com/career/contact-tracer/salaries |
| ***Testing Costs*** | | | | | |
| Hours Spent Per Test Collection & Send Out, PCR | 0.067 | hours | 0 | 0 | BD internal data |
| Hours Spent Per Test Run/Collection, Antigen | 0.05 | hours | 0 | 0 | BD internal data |
| Staff Hourly Wage, University, Running Test/ Test Collection | $25.00 | hours | $13.51 | $45.00 | https://www.cnbc.com/2020/05/20/temperature-screeners-covid-19-testers-these-jobs-were-created-by-pandemic.html |
| ***PPE Costs Per Test*** | | | | | |
| Cost of a Gown | $2.64 | USD | 0 | 0 | https://www.nheconomy.com/getmedia/e1c2a698-e7a1-4e18-8391-b66b52c0bd38/Average_PPE_Price.pdf |
| Cost of a Mask (N95) | $8.68 | USD | 0 | 0 | |
| Cost of Gloves | $0.05 | USD | 0 | 0 | |
| Cost Per Papper | $10.72 | USD | 0 | 0 | |
| Gloves Per Test | $2.00 | USD | 0 | 0 | assumption |
| Masks Per Day of Testing | 2 | unit | 0 | 0 | |
| Gowns per Day of Testing | 2 | unit | 0 | 0 | |
| Pappers Per Day Testing | 1 | unit | 0 | 0 | |
